# Lipid profiles in ST-elevation myocardial infarction with and without diabetes: the gap between prescription and targets

**DOI:** 10.1101/2024.04.15.24305838

**Authors:** Marco Biasin, Nicolò Cordioli, Ilaria Armani, Ludovica Guerrieri, Giulia Parmegian, Alessandro Sarai, Flavio Luciano Ribichini, Alessia Gamabaro

## Abstract

**INTRODUCTION:** Lipids are critical in coronary atherosclerosis, making lipid reduction essential for prevention of cardiovascular disease. Achieving guideline-recommended LDL cholesterol (LDL-C) targets remains challenging for patients with and without diabetes. This study aims to compare clinical differences between STEMI patients with and without diabetes and evaluate lipid-lowering strategies on admission and on discharge.

**METHODS:** Retrospective study on STEMI patients admitted to our center between 2021 and 2023. Data included anthropometric details, lipid profiles, cardiovascular risk scores and drug therapy. Theoretical LLT potency allowed computation of LDL-C levels as if they hadn’t undergone any LLT therapy (wild LDL-C).

**RESULTS:** Of 467 screened patients, 443 were included, with 72 having diabetes. Statistically significant differences were observed in hypertension (72.2% vs. 56.3%, p < 0.001) and peripheral arterial disease prevalence (15.2% vs. 6.2%, p = 0.01). Non-diabetic patients had higher total cholesterol, HDL-C, and LDL-C levels but similar wild LDL-C (110.7 mg/dL vs. 117.5 mg/dL, p = 0.30). At admission, 50.0% (diabetic) and 81.1% (non-diabetic) did not achieve LDL-C targets (p < 0.001). At discharge, 31.9% (diabetic) and 36.6% (non-diabetic) were discharged without sufficient LLT to achieve target cholesterol levels (p < 0.001).

**CONCLUSION:** A significant proportion of patients, especially those with diabetes, failed to attain recommended LDL-C targets at STEMI admission. Aggressive lipid-lowering interventions, in particular with the support of electronic tools to assess LLT potency, are crucial for prompt LDL-C target attainment.

**Highlights:**

- At admission significant proportion of STEMI patients, including those with diabetes, fail to achieve LDL-C targets for their risk class in primary prevention.
- Limited adoption of combination therapies with ezetimibe.
- High-potency statins commonly prescribed at discharge, but many patients forecasted not to achieve LDL-C targets.
- Tailored treatment regimens utilizing electronic tools crucial for prompt attainment of LDL-C targets post-STEMI.

**Graphical abstract:** 

## 1 INTRODUCTION

In recent years, significant progress has been made in the management of lipid profiles for patients undergoing ST-elevation myocardial infarction (STEMI), driven by the evolving landscape of lipid-lowering therapy (LLT) strategies. As lipids play a critical role in the development of coronary atherosclerosis lesions, obtaining a significant reduction of lipid-related risk has long been a crucial aspect of primary and secondary prevention [1].

The presence of diabetes further complicates this scenario, as individuals with both conditions often manifest unique lipid profiles. Indeed, patients with type 2 diabetes mellitus (T2DM) often exhibit an atherogenic lipid profile, increasing their cardiovascular disease risk. Dyslipidemia in T2DM is characterized by elevated triglycerides, decreased HDL cholesterol, and a preponderance of small, dense low-density lipoprotein particles, potentially enhancing atherogenicity through increased susceptibility to oxidation, despite LDL cholesterol (LDL-C) levels similar to non-diabetic individuals [2].

In the sphere of primary prevention, randomized controlled trials with lipid-lowering agents in patients at risk of cardiovascular events (including patients with T2DM) have demonstrated a log-linear proportional reduction of cardiovascular events and mortality for each 1 mmol/L (38,6 mg/dL) reduction of LDL-C [3]. However, many patients (diabetic and non-diabetic) in primary prevention fail to achieve the LDL-C targets recommended by guidelines [4,5]

For patients with a recent myocardial infarction, guidelines recommend achieving an LDL-C level of < 1.4 mmol/L (< 55 mg/dL) with a class 1 recommendation and a level of evidence A [6]. Lipid management, with a primary focus on attaining optimal LDL-C levels, assumes a critical role in post-STEMI care management. It is important not only to achieve the LDL-C targets recommended by guidelines but also to achieve them promptly following a STEMI. This is of paramount importance, as highlighted by the findings of studies that revealed an association between rapid reduction of LDL-C levels and positive clinical outcomes [7,8].

The first purpose of this study is to examine the risk factors, lipid profiles, and lipid-lowering therapies on admission in patients presenting with STEMI. The focus will be kept on finding different characteristics of patients affected by diabetes and patients free from this condition.

The second objective of this investigation is to assess the effectiveness of lipid-lowering management strategies implemented at the point of hospital discharge. The aim is to ascertain the alignment of these strategies with guideline-recommended target lipid level.

## 2 METHODS

This descriptive retrospective study was conducted utilizing patient data between January 2021 and December 2023 extracted from the archives of our Cardiology division, a public large university hospital in Europe. Inclusion criteria were: patients admitted with a confirmed diagnosis of STEMI, having a complete lipid profile on lab analysis and a reliable pharmacological and clinical history on admission. Exclusion criterion from the study was the patient’s death during index hospitalization.

Collected data encompassed anthropometric and demographic details, lipid profiles, cardiovascular risk factors, the presence of chronic coronary disease, prior stroke, drug therapy on admission and discharge. We consider high-potency statin: Rosuvastatin 40 or 20 mg and Atorvastatin 80 or 40 mg; the others were considered low-potency statin.

The lipid parameters examined included total cholesterol (tChol), LDL-C calculated using both Friedwald (F-LDL-C) and Hopkins Martin (M-LDL-C) formulas, high-density lipoprotein cholesterol (HDL-C) and triglycerides (TG). By utilizing theoretical lipid lowering therapy potency derived by meta-analysis on cholesterol trials, LDL levels were also computed for all patients as if they hadn’t undergone any LLT (wild LDL-C) [9]. In order to perform our analysis, we employed an electronic tool provided by the Italian Association of Hospital Cardiologists (ANMCO) called “LDL-C Goal”, this tool offers a comprehensive framework for calculating lipid profiles, including LDL-C levels, considering factors such as baseline lipid panel and baseline LLT [10]. Using this tool and knowing the LDL-C levels achievable by the patient after the discharge were calculated.

We evaluated the CVD risk of patients upon admission, utilizing available medical record data and applying ESC CVD risk scores (SCORE2, SCORE2-OP, SCORE2-Diabetes) [11]. Using the risk class derived from the scores, we determined the LDL-C targets the patient would have reached prior to the STEMI event [12]. The patients’ risk class at discharge was categorized as very high (with a target LDL-C <55) or extremely high risk (if patients experienced a second vascular event within 2 years), corresponding to a target LDL-C of < 40 mg/dL.

Ethical approval was secured from the institutional review board, and the study adhered to the principles of patient confidentiality.

Continuous variables that did not deviate substantially from the normal distribution were reported as mean and standard deviation; otherwise, they were reported as median and interquartile range. Categorical variables were reported as frequencies and percentages. Associations were calculated using the Mann-Whitney U Test. A two-sided p Value of < 0.05 was considered to be statistically significant. All statistical analyses were performed by using SAS version 9.4 (SAS Institute, Inc., Cary, North Carolina).

## 3 RESULTS

### 3.1 Demographic variables and clinical characteristics

Of the 467 patient records screened we excluded 24 patients (2 turned out to not been STEMI, 12 dead during index hospitalization and 9 had incomplete data). The remaining 443 participants fulfilled the inclusion criteria. Among them, 72 patients had diabetes (2 with type 1 diabetes and 70 with type 2). The majority of participants were male in both diabetic and non-diabetic cohorts (79.1% [57/72] vs 79.8% [296/371], p = 0.93) and the average age of the patients was also similar between the two cohorts (65.1±11.9 vs 64.9±13.5, p = 0.92).

The clinical characteristics of the two cohorts are summarized in Table 1. The prevalence of risk factors exhibited statistical differences between diabetic and non-diabetic cohorts only for hypertension, which was more frequent among diabetic patients (72.2% [52/72] vs. 56.3% [209/371], p < 0.001), and peripheral arterial disease, which was also more frequent in diabetic patients (15.2% [11/72] vs. 6.2% [23/371], p = 0.01).

**Table 1.** Demographic and clinical characteristics.

|  | **Diabetic**<br>**(n = 72)** | **Non diabetic**<br>**(n = 371)** | **p Value** |
| --- | --- | --- | --- |
| **Age, yrs** | 65.1±11.9 | 64.9±13.5 | 0.92 |
| **Male** | 57 (79.1%) | 296 (79.8%) | 0.93 |
| **BMI** | 26.4± 4.5 | 26.3± 4.0 | 0.89 |
| **Hypertension** | 52 (72.2%) | 209 (56.3%) | 0.016 |
| **Active smokers** | 24 (33.3%) | 152 (40.9%) | 0.28 |
| **Family history of CVD** | 21 (29.1 %) | 106 (28.5 %) | 0.97 |
| **eGFR < 30 ml/min/1.73m2** | 0 (0.0%) | 6 (1.6%) | 0.70 |
| **Peripheral arterial disease** | 11 (15.2%) | 23 (6.2 %) | 0.015 |
| **History of stroke** | 3 (4.2%) | 8 (2.1%) | 0.83 |

### 3.2 Baseline Lipid profile and Lipid-lowering therapy

Statistically significant differences were found in the lipid profiles of the two cohorts. Specifically, tChol, HDL-C and LDL-C were higher in non-diabetic patients compared to diabetic patients, while TG were higher among diabetic patients (Table 2). Compared to LDL-C, the wild LDL-C values showed a less pronounced difference between diabetic and non-diabetic patients using both the Friedwald formula (105.6 mg/dL [IQR 60.2-137.9] vs. 116.2 mg/dL [IQR 88.2-136.3], p = 0.10) and the Martin formula (110.7 mg/dL [IQR 66.8-136.6] vs. 117.5 mg/dL [IQR 91.1-138.0], p = 0.30).

**Table 2.**
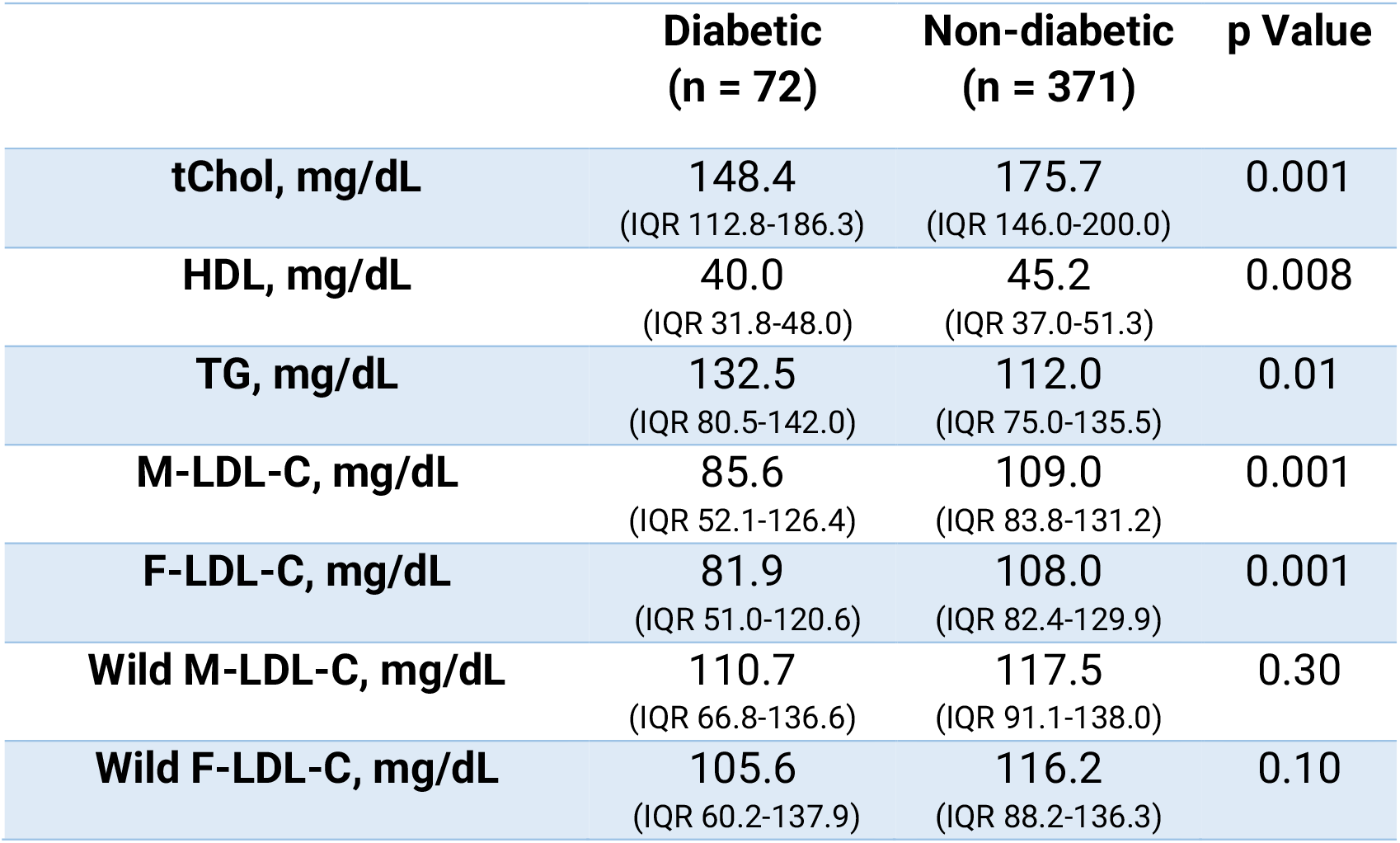
Baseline lipid profile.

Tabel 2 Baseline lipid profile
|  | **Diabetic**<br>**(n = 72)** | **Non-diabetic**<br>**(n = 371)** | **p Value** |
| --- | --- | --- | --- |
| **tChol, mg/dL** | 148.4<br>(IQR 112.8-186.3) | 175.7<br>(IQR 146.0-200.0) | 0.001 |
| **HDL, mg/dL** | 40.0<br>(IQR 31.8-48.0) | 45.2<br>(IQR 37.0-51.3) | 0.008 |
| **TG, mg/dL** | 132.5<br>(IQR 80.5-142.0) | 112.0<br>(IQR 75.0-135.5) | 0.01 |
| **M-LDL-C, mg/dL** | 85.6<br>(IQR 52.1-126.4) | 109.0<br>(IQR 83.8-131.2) | 0.001 |
| **F-LDL-C, mg/dL** | 81.9<br>(IQR 51.0-120.6) | 108.0<br>(IQR 82.4-129.9) | 0.001 |
| **Wild M-LDL-C, mg/dL** | 110.7<br>(IQR 66.8-136.6) | 117.5<br>(IQR 91.1-138.0) | 0.30 |
| **Wild F-LDL-C, mg/dL** | 105.6<br>(IQR 60.2-137.9) | 116.2<br>(IQR 88.2-136.3) | 0.10 |

Diabetic patients demonstrate a statistically significant higher prevalence of LLT on board (51.4% [37/72] vs 11.0% [41/371], p <0.001). The majority of patients on statin therapy had a low-intensity statin on board, regardless of whether they were affected by diabetes or not (78.3% [29/37] vs. 70.7% [29/41], p = 0.60).

Among patients receiving statins, 9.7% (7/72) of diabetic patients and 3.5% (13/371) of non-diabetic patients received a combination therapy with ezetimibe (p = 0.04). Only 1 patient with diabetes was on therapy with PCSK9 inhibitor (PCSK9i). The average LDL-C reduction potency of the LLT onboard at the time of admission was 43.2 ± 11.1% in diabetic patients versus 42.3 ±12.1% in non-diabetic patients (p = 0.89) (Table 3).

**Table 3.**
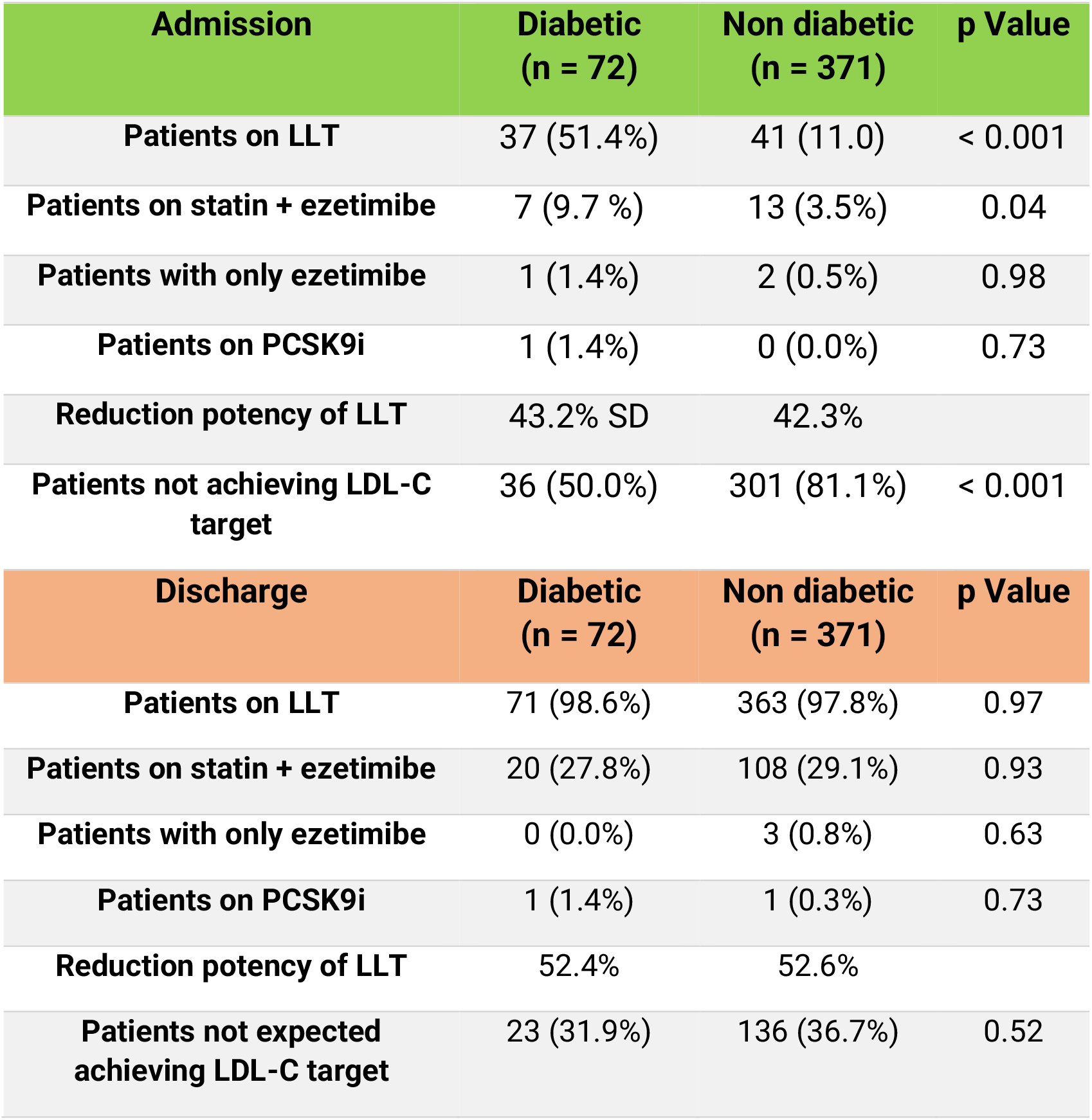
Strategies of the lipid-lowering therapy at hospital admission and discharge.

Tabel 3 Strategies of the lipid-lowering therapy at hospital admission and discharge
| Admission | Diabetic<br>(n = 72) | Non diabetic<br>(n = 371) | p Value |
| --- | --- | --- | --- |
| Patients on LLT | 37 (51.4%) | 41 (11.0) | < 0.001 |
| Patients on statin + ezetimibe | 7 (9.7 %) | 13 (3.5%) | 0.04 |
| Patients with only ezetimibe | 1 (1.4%) | 2 (0.5%) | 0.98 |
| Patients on PCSK9i | 1 (1.4%) | 0 (0.0%) | 0.73 |
| Reduction potency of LLT | 43.2% SD | 42.3% |  |
| Patients not achieving LDL-C target | 36 (50.0%) | 301 (81.1%) | < 0.001 |
| Discharge | Diabetic<br>(n = 72) | Non diabetic<br>(n = 371) | p Value |
| Patients on LLT | 71 (98.6%) | 363 (97.8%) | 0.97 |
| Patients on statin + ezetimibe | 20 (27.8%) | 108 (29.1%) | 0.93 |
| Patients with only ezetimibe | 0 (0.0%) | 3 (0.8%) | 0.63 |
| Patients on PCSK9i | 1 (1.4%) | 1 (0.3%) | 0.73 |
| Reduction potency of LLT | 52.4% | 52.6% |  |
| Patients not expected achieving LDL-C target | 23 (31.9%) | 136 (36.7%) | 0.52 |

### 3.3 Baseline CVD risk score and patients on lipid targets at hospital admission

We evaluated the CVD risk score in patients at the moment of admission. In the diabetic cohort, 54.2% (39/72) were at very high risk, 27.8% (20/72) were at high risk, and 18.1% (13/72) were at moderate or low risk. Among non-diabetic patients 21.0% (78/371) were at very high risk, 51.2% (190/371) were at high risk, and 27.8% (103/371) were at moderate or low risk (Figure 1). The percentage of patients not achieving the recommended LDL-C targets for their risk class was 50.0% (36/72) in the diabetic cohort and 81.1% (301/371) in the non-diabetic cohort (p < 0.001) (Figure 2).

**Figure 1.**
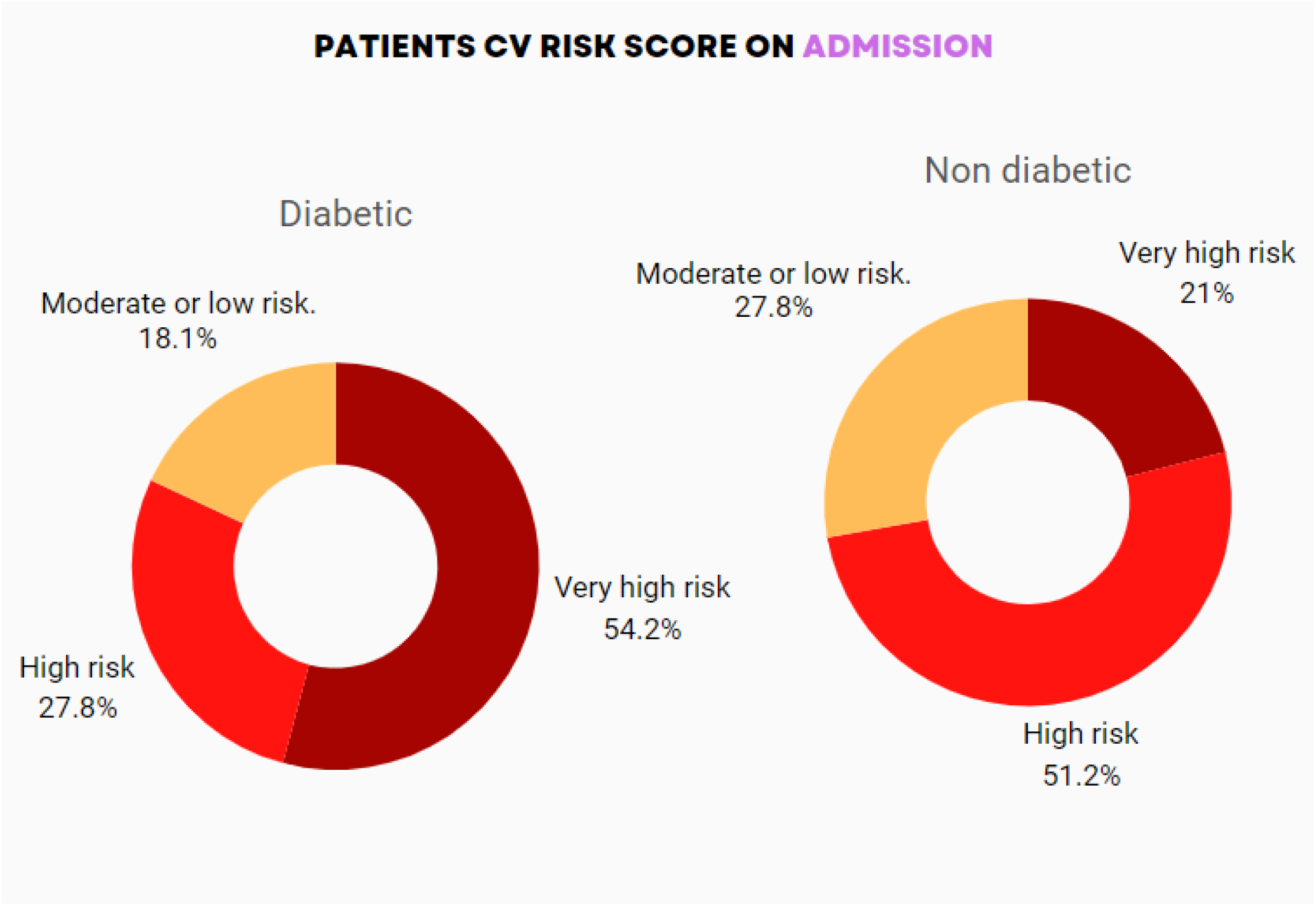
Patients CVD risk score on admission

**Figure 2.**
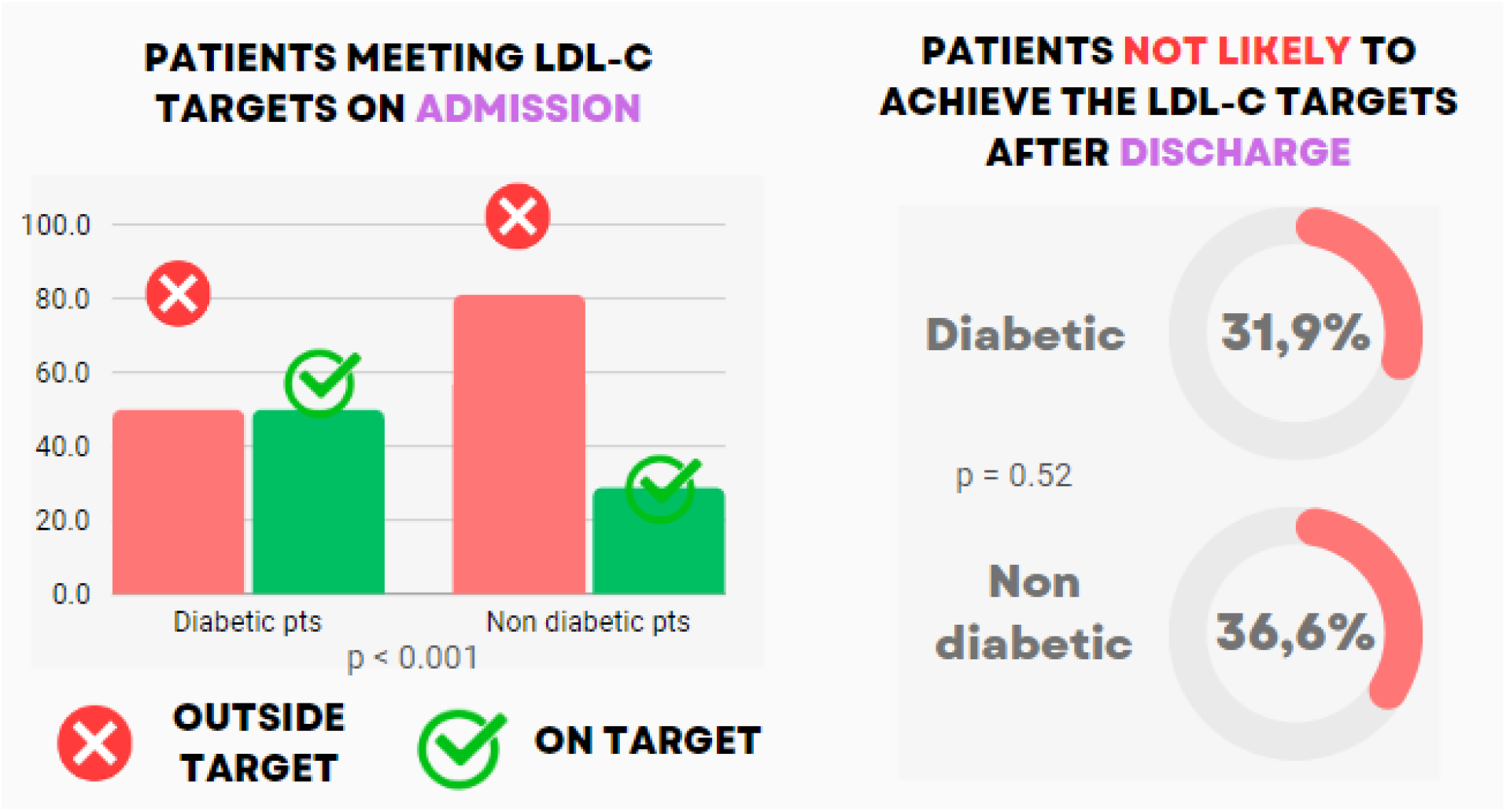
Patients meeting LDL-C targets on hospital admission and discharge

### 3.4 Management strategies of the lipid-lowering therapy at hospital discharge

At the time of discharge, 439 out of 443 patients were classified as very high risk, with 4 out of 443 classified as extremely high risk. Out of the 443 discharged patients, 93.9% (416/443) received high-potency statins, 4.3% (19/443) were prescribed low-potency statins and 1.8% (8/443) of patients were prescribed without statins. Among patients receiving high-potency statins, 28.8% (120/416) received a combination with ezetimibe, and 0.4% (2/416) received a combination with both ezetimibe and PCSK9 inhibitor (PCSK9i). LLT was similar in both diabetic and non-diabetic cohorts (Table 3).

The average LDL reduction potency of the LLT prescribed at discharge was comparable between diabetic and non-diabetic patients (52.4 ± 7.6 % vs. 52.6 ± 8.1 %, p = 0.42). The rate for combination therapy with high-potency statins and ezetimibe observed in the two cohorts was 27.8% (20/72) among diabetic patients and 29.1% (108/371) among non-diabetic patients (p= 0.93).

At the time of discharge, 35.8% (159/443) of patients were discharged with a LLT that was not expected to be sufficient to enable them to achieve cholesterol targets for their risk class. This was true for both diabetic and non-diabetic patients (31.9% (23/72) vs. 36.6% (136/371), p = 0.52) (Figure 2). Patients who did not reach the LDL-C target as per guidelines had, on average, a value of 16.0 ± 22.9 mg/dL higher than the target value. In the diabetic group, this value was higher by 21.9 ± 21.7 mg/dL, while in non-diabetics it was 14.1 ± 20.2 mg/dL (p = 0.01). If every patient had been discharged with the highest combinational therapy available (Rosuvastatin plus Ezetimibe 40/10 mg) only 6.9% (5/72) diabetic would not have reached the LDL-C target as per guidelines, and 3.2% (12/371) of non-diabetic patients.

## 4 DISCUSSION

Demographic characteristics between the two cohorts were largely similar; however, notable differences emerged in clinical features, particularly in the prevalence of hypertension and peripheral arterial disease, which were more prevalent among diabetic patients. These findings are consistent with existing literature [13,14] and partly likely contribute to explaining the more elevated level of CVD risk observed in diabetic patients.

While lipid profiles appeared different between diabetic and non-diabetic patients, with diabetic patients showing lower levels (except for TG), it’s essential to interpret these results in the context of baseline statin use. The higher prevalence of statin therapy among diabetic patients likely influences lipid profiles, as indicated by the comparable levels of wild LDL-C between the two groups. This finding aligns with the statement made in the introduction, indicating that dyslipidemia in T2DM is characterized by elevated TG, decreased HDL-C despite LDL-C levels similar to non-diabetic individuals [2].

More than half of diabetic patients were classified as very high CVD risk, while in non-diabetic patients, the proportion of patients at very high CVD risk was lower. Nevertheless, over two-thirds of patients across both cohorts were deemed at high or very high CVD risk. Despite this, a significant proportion of patients, regardless of diabetic status, failed to attain LDL-C targets [4,15,16].

It should be emphasized that to assess the LDL-C target, we referred to the Martin formula because, in a large cross-sectional analysis, this method of LDL-C estimation was found to be more accurate than the Friedewald equation for both fasting and non-fasting. Specifically, the Martin formula outperformed the Friedewald equation when LDL-C was <70 mg/dL or when TG levels were high. [17]

The low number of patients reaching lipid targets upon admission underscores a gap in primary prevention efforts, potentially resulting from underutilization of LLT, particularly among non-diabetic patients, widespread utilization of low-intensity statins in primary prevention, and the limited adoption of statin-ezetimibe combinations. This observation is critical because the inadequate utilization of LLT in primary prevention fail to adequately reduce cardiovascular risk and potentially leading to avoidable CV events [18].

The use of ezetimibe in combination with statins is important for two reasons. First, ezetimibe added to statin therapy results in an additional 15–20% reduction in LDL-C cholesterol. In fact, the addition of ezetimibe to statin therapy has been shown to be significantly more effective in achieving target goals than doubling the statin monotherapy dose. The second reason is the presence of statin muscle-related adverse events (SAMS), which limit the use of statins. Although the true incidence of SAMS is uncertain, occurring in 1.5–5% in randomized controlled trials, this percentage increases in observational studies, reaching 10–33%[19]. Ezetimibe allows for increasing the effectiveness of therapy without the need for higher doses, thereby reducing the risk of statin-associated muscle symptoms SAMS [20].

Notably, diabetic patients may receive more statins maybe because they are perceived with higher CVD risk, whereas this risk may not be accurately assessed and managed in non-diabetic patients, as reflected by only 11.1% of non-diabetic patients meeting lipid targets for their risk class at STEMI admission. This data is consistent with other population-based observational studies that show low usage of statins in primary prevention [5].

At discharge, high-potency statins were prescribed to nearly all patients with an average LLT potency exceeding 50%. However, it’s noteworthy that while the use of combination therapies with ezetimibe is higher than in primary prevention, it is prescribed to less than a third of the patients. Similarly, the use of PCSK9 inhibitors remained scarce, partly due to reimbursement constraints in the Italian healthcare system, hindering their implementation during STEMI hospitalization. Consequently, over a third of patients were forecasted not to achieve target LDL-C levels, with an average LDL-C deviation from target of 16 ± 22.9 mg/dL. This underscores the critical need for more effective lipid-lowering interventions, even during hospitalization, to expedite the attainment of LDL-C target levels.

To achieve this, combination therapies of high-intensity statins with ezetimibe are undoubtedly beneficial. In fact, a recent multicenter analysis demonstrated that upfront combination LLT is superior to statin monotherapy in reducing all-cause mortality [21]. This challenges the stepwise approach and confirms the advantage of utilizing upfront combination strategies, particularly in patients with very high CVD risk.

It is noteworthy that conducting a simulation where all patients analyzed in our study were discharged with the most effective combination therapy available on the market (Rosuvastatin/Ezetimibe 40/10 mg) reveals that over 90% of patients would have been able to achieve their LDL-C target. However, it is important to recognize that this therapy cannot be prescribed to all patients due to the higher incidence of SAMS and the contraindication in patients with chronic kidney disease.

The current ESC/EAS guidelines suggest re-evaluating LDL-C levels 4–6 weeks after ACS to determine whether initiation of ezetimibe or PCSK-9 inhibitors is necessary. However, delaying intervention until the first follow-up visit risks prolonging the time to achieve guideline-recommended LDL-C levels. Therefore, tailoring treatment regimens based on individual patient baseline lipid profiles, utilizing electronic tools to assess whether the prescribed LLT potency at discharge is adequate to reach the desired LDL-C target in the shortest possible time is crucial as delayed intervention may worsen outcomes.

### 4.1 LIMITATIONS

Several limitations must be acknowledged in this study. Firstly, the retrospective design introduces inherent biases, limiting the establishment of causality and generalizability of findings. Secondly, the study was conducted at a single center, which may not fully represent the diversity of patient populations and treatment practices in other healthcare settings. Thirdly, the study focused solely on lipid profiles and lipid-lowering therapies, excluding other potential confounders or variables that could impact clinical outcomes. Lastly, the study did not assess long-term follow-up data, which could provide valuable insights into the durability and sustainability of lipid-lowering interventions beyond the acute hospitalization period.

## 5 CONCLUSIONS

In conclusion, our study highlights several key findings regarding lipid profiles and LLT management in patients with STEMI. Despite advances in LLT strategies, a significant proportion of patients, both diabetic and non-diabetic, failed to achieve guideline-recommended LDL-C targets, indicating gaps in primary prevention efforts. The use of high-potency statins was common at discharge, but there was limited adoption of combination therapies with ezetimibe, which could potentially optimize LDL-C management. The study underscores the importance of tailored treatment regimens utilizing electronic tools to assess the adequacy of prescribed LLT potency at discharge based on baseline lipid profile and baseline LLT to expedite the attainment of LDL-C targets after a STEMI.

## Data Availability

All data produced in the present work are contained in the manuscript

